# COVID Symptoms, Symptom Clusters, and Predictors for Becoming a Long-Hauler: Looking for Clarity in the Haze of the Pandemic

**DOI:** 10.1101/2021.03.03.21252086

**Authors:** Yong Huang, Melissa D. Pinto, Jessica L. Borelli, Milad Asgari Mehrabadi, Heather Abrihim, Nikil Dutt, Natalie Lambert, Erika L. Nurmi, Rana Chakraborty, Amir M. Rahmani, Charles A. Downs

## Abstract

Emerging data suggest that the effects of infection with SARS-CoV-2 are far reaching extending beyond those with severe acute disease. Specifically, the presence of persistent symptoms after apparent resolution from COVID-19 have frequently been reported throughout the pandemic by individuals labeled as “long-haulers”. The purpose of this study was to assess for symptoms at days 0-10 and 61+ among subjects with PCR-confirmed SARS-CoV-2 infection. The University of California COvid Research Data Set (UC CORDS) was used to identify 1407 records that met inclusion criteria. Symptoms attributable to COVID-19 were extracted from the electronic health record. Symptoms reported over the previous year prior to COVID-19 were excluded, using nonnegative matrix factorization (NMF) followed by graph lasso to assess relationships between symptoms. A model was developed predictive for becoming a long-hauler based on symptoms. 27% reported persistent symptoms after 60 days. Women were more likely to become long-haulers, and all age groups were represented with those aged 50 ± 20 years comprising 72% of cases. Presenting symptoms included palpitations, chronic rhinitis, dysgeusia, chills, insomnia, hyperhidrosis, anxiety, sore throat, and headache among others. We identified 5 symptom clusters at day 61+: chest pain-cough, dyspnea-cough, anxiety-tachycardia, abdominal pain-nausea, and low back pain-joint pain. Long-haulers represent a very significant public health concern, and there are no guidelines to address their diagnosis and management. Additional studies are urgently needed that focus on the physical, mental, and emotional impact of long-term COVID-19 survivors who become long-haulers.

## Introduction

In the United States, over 28 million people have been infected with SARS-CoV-2, the virus responsible for COVID-19 (1), and the cumulative hospitalization rate has exceeded 1300 persons per 100,000 since early 2020 (2). Hospitalized patients account for 1% of COVID-19 patients, yet most research to-date has focused on in-patients with severe disease. However, very little is known about the medium-and long-term consequences of COVID-19 among non-hospitalized individuals, although emerging data suggest a significant proportion of these subjects experience persistent symptoms associated with antecedent SARS-CoV-2 infection. Those with persistent symptoms have been labeled “long-haulers” or persons with long COVID-19. Recent estimates suggest that ∼10% of hospitalized patients go on to become long-haulers (3-5). However, the body of evidence regarding long-haulers, particularly among the 99% of non-hospitalized cases, is nascent.

Late sequelae following an infectious disease is not uncommon. However, it is unclear whether clinical manifestations reflect primary organ involvement during an acute infection or if long-term signs and symptoms are promoted by aberrant inflammatory immune response. Understanding the late sequelae of SARS-CoV-2 infection is limited due to small sample sizes and a preponderance of studies that have focused on hospitalized survivors, with very limited data at the population level. Further studies examining long-term outcomes in subjects with “milder” infection are crucial to understanding both the pathophysiology and the public health impact of COVID-19. In addition, developing an understanding of host factors that predict long-hauler status as well as potential association with symptom clusters will be pivotal to the development of evidence-based management guidelines.

In the current study, we utilized electronic health records (EHR) from community dwelling individuals (*N*=1407) with confirmed SARS-CoV-2 infection (via PCR) to determine symptoms and symptom clusters. Specifically, we evaluated symptoms at presentation (days 0-10 following a COVID-19 diagnosis) and at days 61+. We defined long-haulers as persons with persistent symptoms at day 61+ (27%) and evaluated if early symptoms or non-modifiable factors (age, ethnicity) could predict likelihood of persistent symptoms at day 61+ (e.g. long-hauler) and/or assignment within any given symptom cluster.

## Methods

### Sample Size and Inclusion Criteria

University of California COvid Research Data Set ((UC CORDS) as of 02/04/2021) contains deidentified electronic records for 178971 patients treated in the state of California. Strict study eligibility criteria were used to obtain a sample of never-hospitalized SARS-CoV-2 infected individuals with COVID-19 symptoms. Patients hospitalized for COVID-19 were excluded. Other inclusion criteria were individuals with at least a 5 year history within the University of California (UC) system, a confirmed positive RT-PCR test for SARS-CoV-2, and only symptoms attributable to SARS-CoV-2 were included as symptoms that were reported within the year prior to testing positive for SARS-CoV-2 were excluded from the analysis, records of healthcare interaction following COVID-19 diagnosis, and individuals with reinfection. A total of 1407 participants met criteria.

### Non-Negative Matrix Factorization for Subgroup Identification

Nonnegative matrix factorization (NMF) extracts sparse and meaningful features from nonnegative data from high dimensional data sets (6) and has been widely used in various domains including image processing, data mining and genomics to extract interpretable components from data. Given a binary symptom occurrence matrix Xn*s, where n is the number of patients, s is the number of symptoms of interest and each entry denotes the occurrence of a symptom for a specific patient. NMF finds two non-negative matrices (W, H) whose product approximates the original non-negative matrix X. W is a low-rank matrix with the rank being the number of components (subgroups) and the entries at each row of W matrix represent how likely a patient falls into each component. The columns of the H matrix illustrate the strength of associations between a component and symptoms, i.e. which symptoms are dominant within a subgroup and which symptoms are less prevalent. Here, NMF was used to extract reported symptoms from the EHR, and then used to identify subgroups of COVID-19 patients in order to identify dominant symptoms within each subgroup (symptom clusters).

### Network Analysis on Symptom Associations

Following NMF we used graphical lasso (glasso) to determine relationships between symptoms at each stage (7). Glasso operates on symptom counts data and yields pairwise associations of symptoms which can be visualized in a network. After network structure estimation using glasso, walktrap algorithm was employed to cluster nodes in the network (8).

### Long-Hauler Predictor Analysis

In order to identify factors leading to the development of persistent symptoms we developed a predictive model that inputs multiple potential key factors to predict if a subject with SARS-CoV-2 infection will become a long-hauler. By inspecting the model’s coefficient strength, we can identify the key predictors. We formulated socio demographics, symptoms the individuals experienced in the first 11 days and if a patient was asymptomatic in the first 11 days as input features and then applied logistic regression as our predictive model. One benefit of this method is that we could not only identify predictors that are associated with higher risk of becoming a long-hauler, but we were also able to identify predictors that may protect a subject from becoming a long-hauler. To alleviate the effect of multicollinearity which may cause misinterpretation of predictor importance, we performed hierarchical clustering on the Spearman rank-order correlations of input features.

## Results

### Features and Symptoms Among Community Dwelling Individuals with COVID-19: Days 0-10 and 61+

Table 1 shows sample distribution related to age, ethnicity, and sex. Figure 1 shows distribution of individuals reporting symptoms at days 0-10; approximately 68% of the total group exhibited symptoms, with 32% being asymptomatic. Prevalent symptoms during this time include (in descending order) dyspnea, cough, fever, chest pain, diarrhea, anxiety, and fatigue (See Figure 1B). Using NMF five symptom clusters with the co-occurrence of symptoms were identified. Symptom network analysis was used to identify prominent symptoms (larger node equates to greater prominence) and the strength of their association with other symptoms, wherein the darker the line connecting nodes indicates a stronger relationship (See Figure 1C).

**Table 1.** Study Demographics

| Socio demographics | Number of subjects | % of subjects |
| --- | --- | --- |
| **Age** |  |  |
| < 18 | 34 | 2% |
| 18-29 | 186 | 13% |
| 30-39 | 232 | 16% |
| 40-49 | 253 | 18% |
| 50-59 | 273 | 19% |
| 60-69 | 225 | 16% |
| 70-79 | 126 | 9% |
| >= 80 | 78 | 6% |
| **Ethnicity** |  |  |
| White | 370 | 26% |
| Black | 51 | 4% |
| Asian | 78 | 6% |
| Hispanic | 766 | 54% |
| Other | 142 | 10% |
| **Sex** |  |  |
| Male | 653 | 46% |
| Female | 754 | 54% |
| **Symptom development** |  |  |
| Long hauler | 382 | 27% |
| Total | 1407 | 100% |

**Figure 1.**
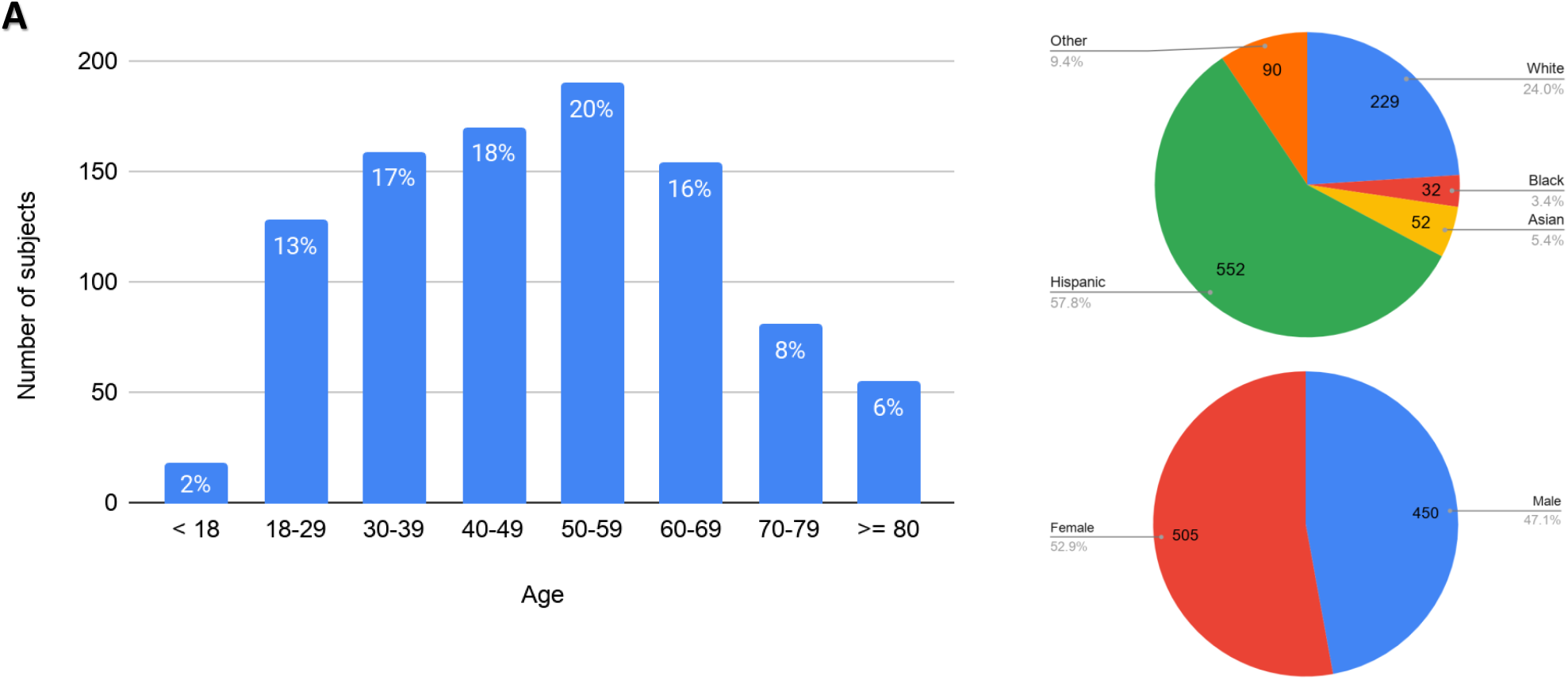
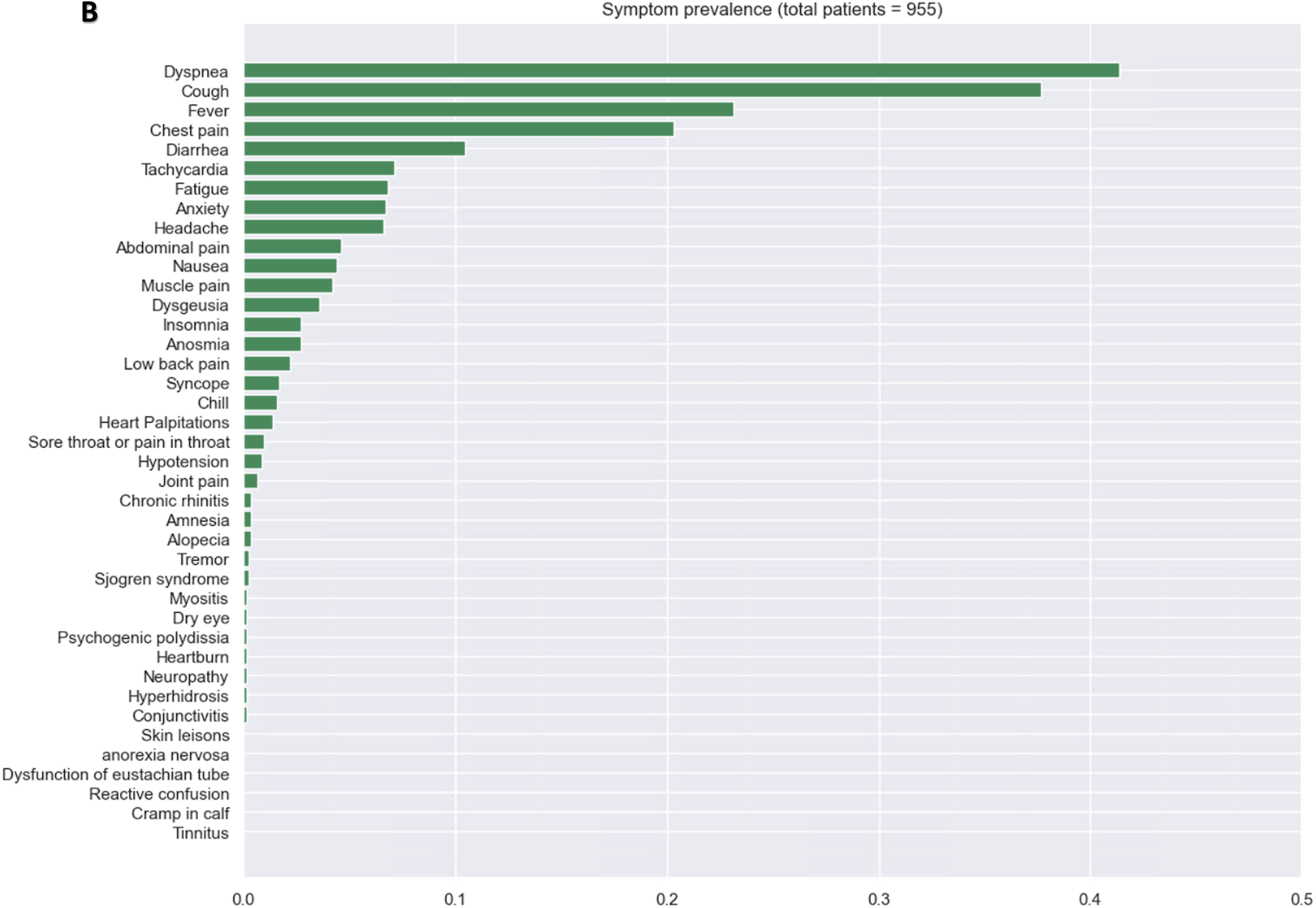
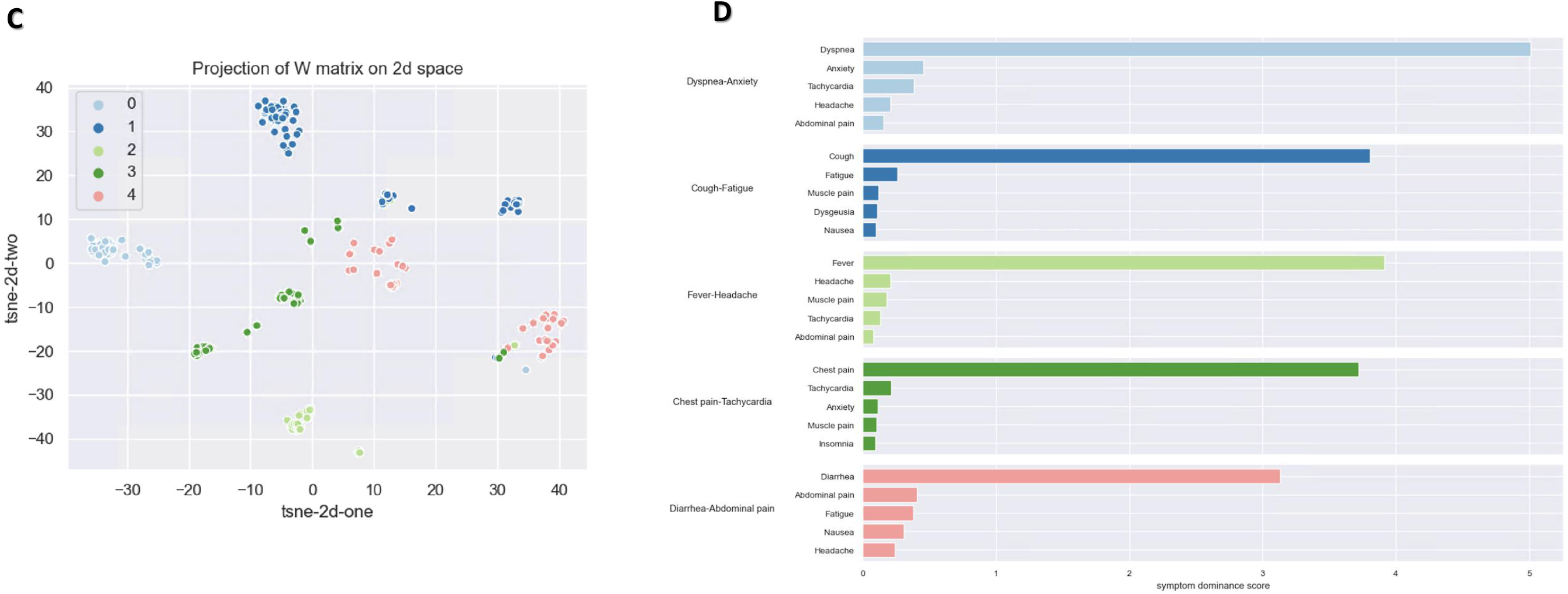
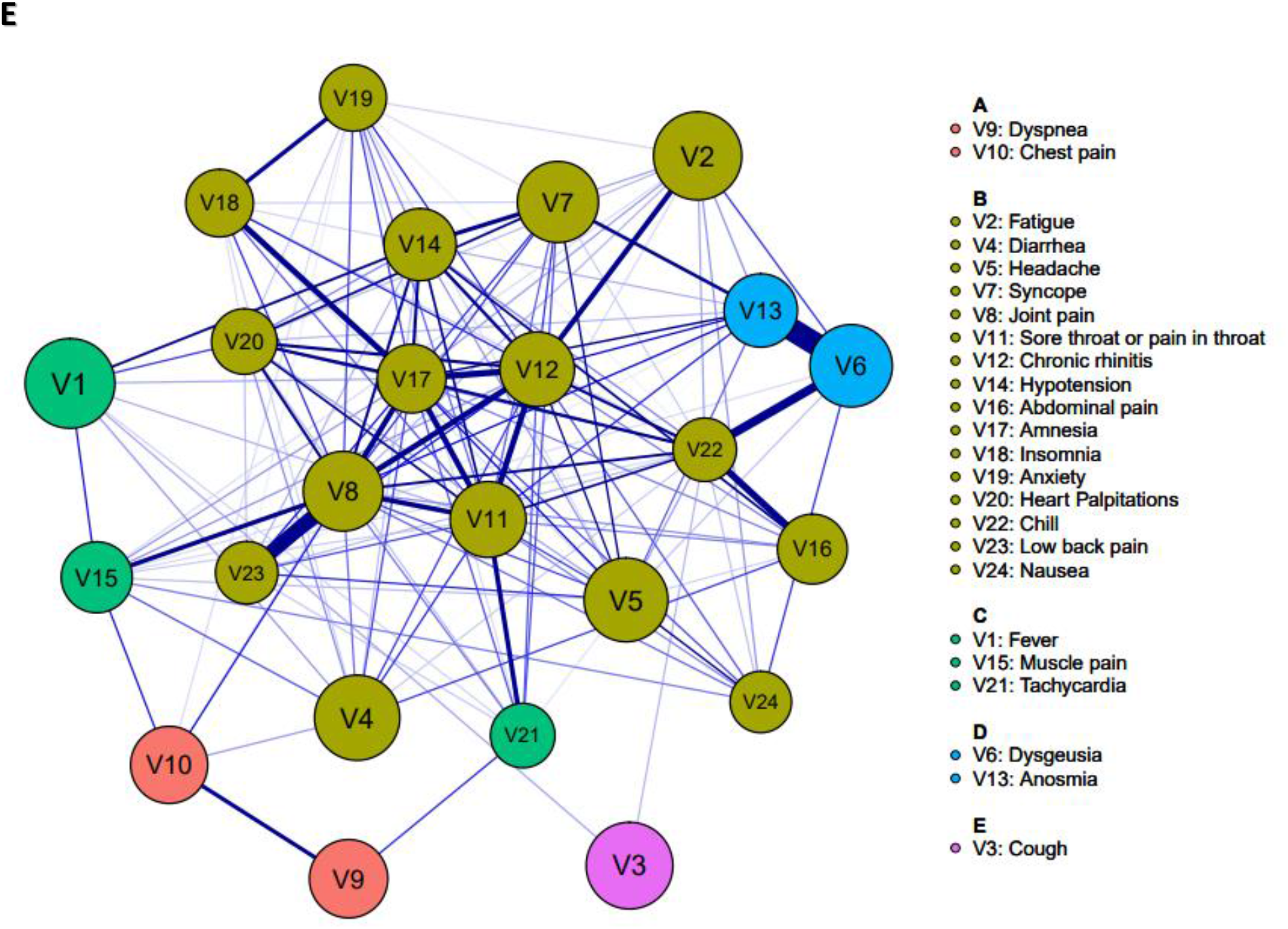
Demographics and symptoms prevalence among SARS-CoV-2 infected community dwellers at days 0-10. (A) Bar and pie graphs showing distribution of age, ethnicity, and sex demographics (N=1345) at days 0-10. (B) Bar graph showing prevalence of symptoms reported at days 0-10. (C) Graph of symptom clusters with (D) corresponding bar graphs demonstrating symptom ranking within each cluster, and (E) symptom network analysis showing relationship between each reported symptom. Each symptom is denoted as a node and the darker the line connecting symptoms indicates a stronger relationship.

Figure 2 depicts the non-modifiable characteristics of long-haulers reporting symptoms at 61+ days. This group was distributed across all age-groups, including among those ≤ 18 (34 participants, a mean age of 9.29 years, 11/34 were ≤ age 5), ethnicities, and included more women than men. Symptoms that are prevalent among long-haulers include (descending order): chest pain, dyspnea, anxiety, abdominal pain, cough, low back pain, and fatigue (See Figure 2B). Using NMF five discrete clusters were identified and symptom network analysis identified prominent symptoms and the strength of their association with other symptoms.

**Figure 2.**
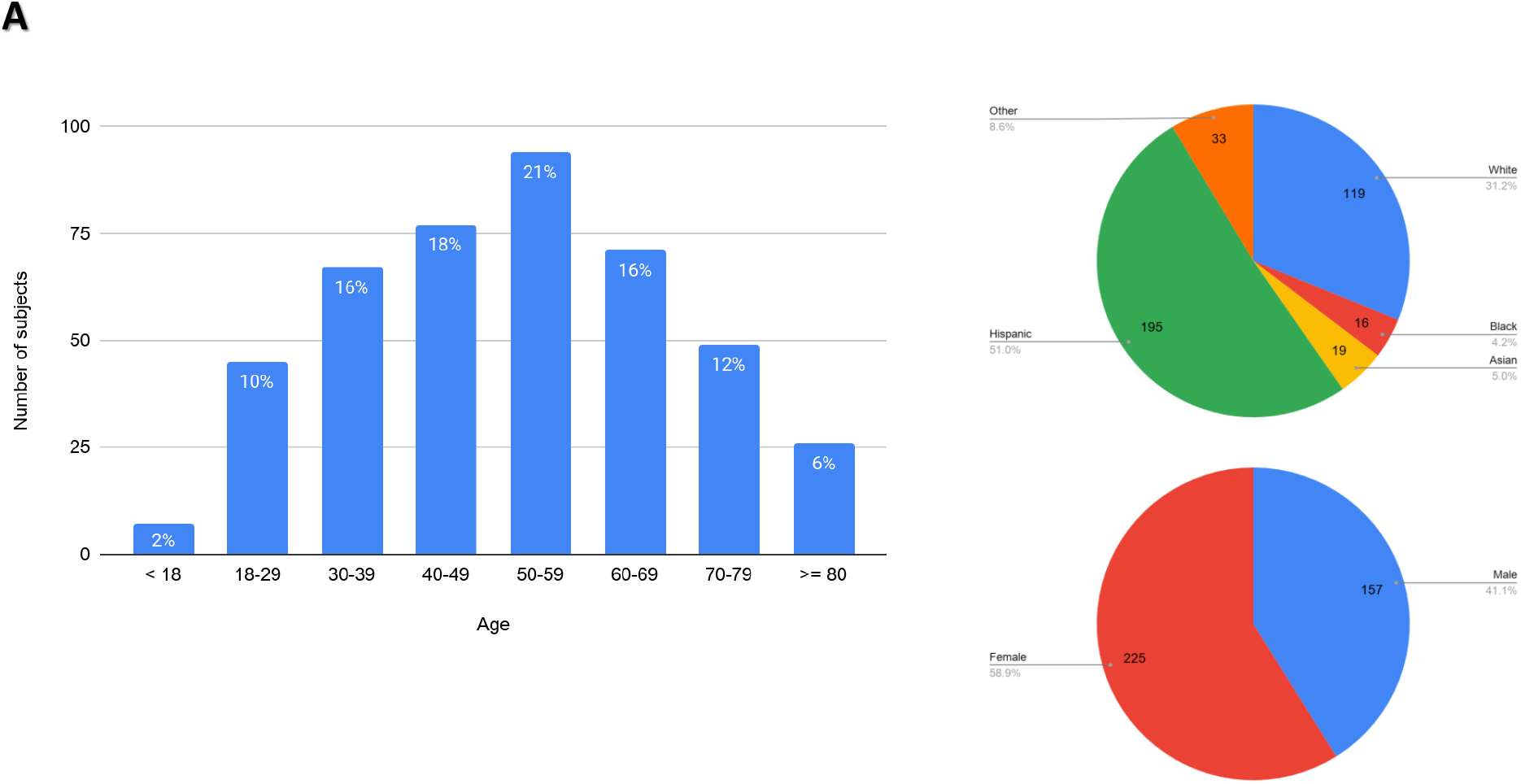
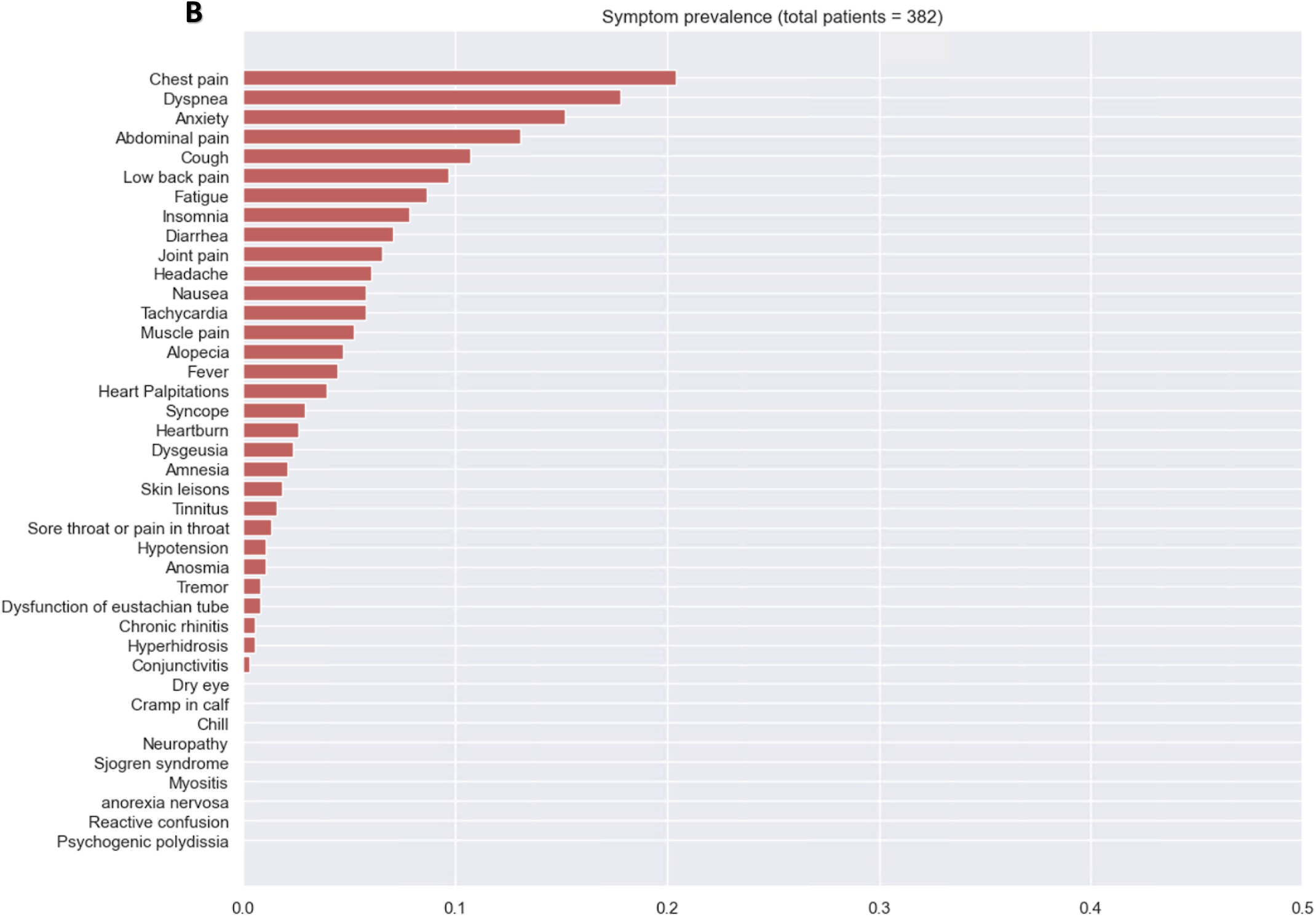
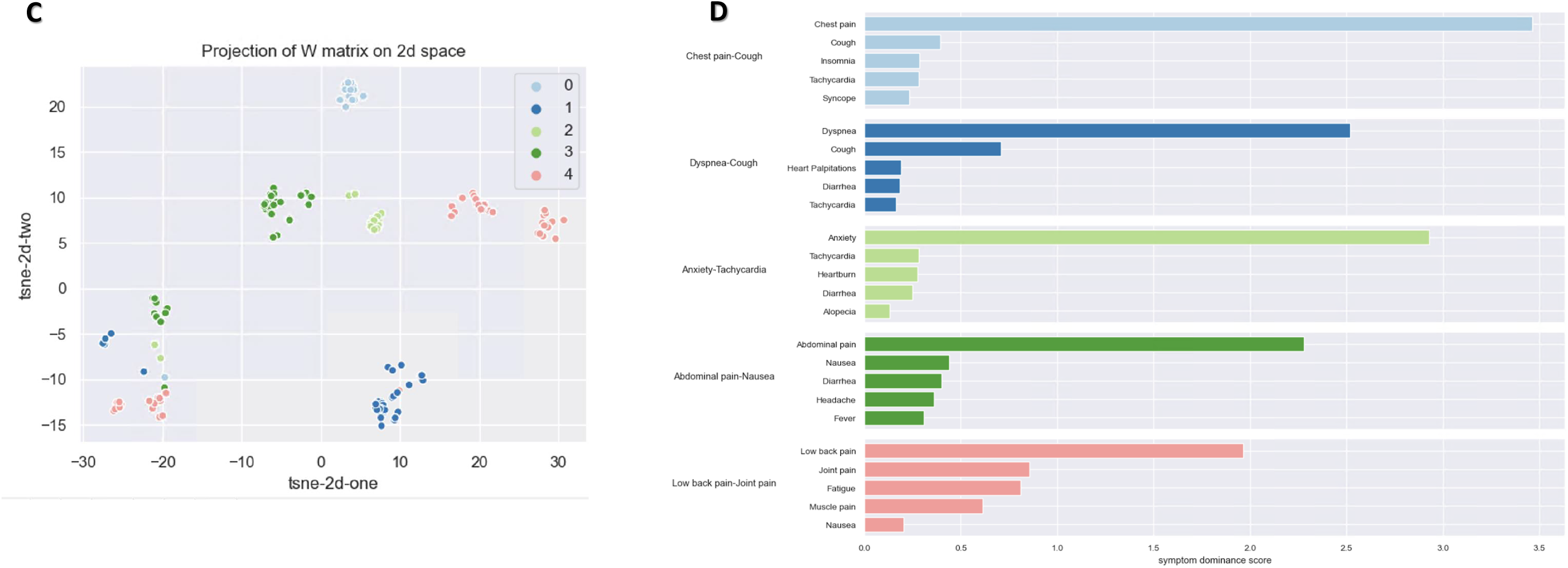
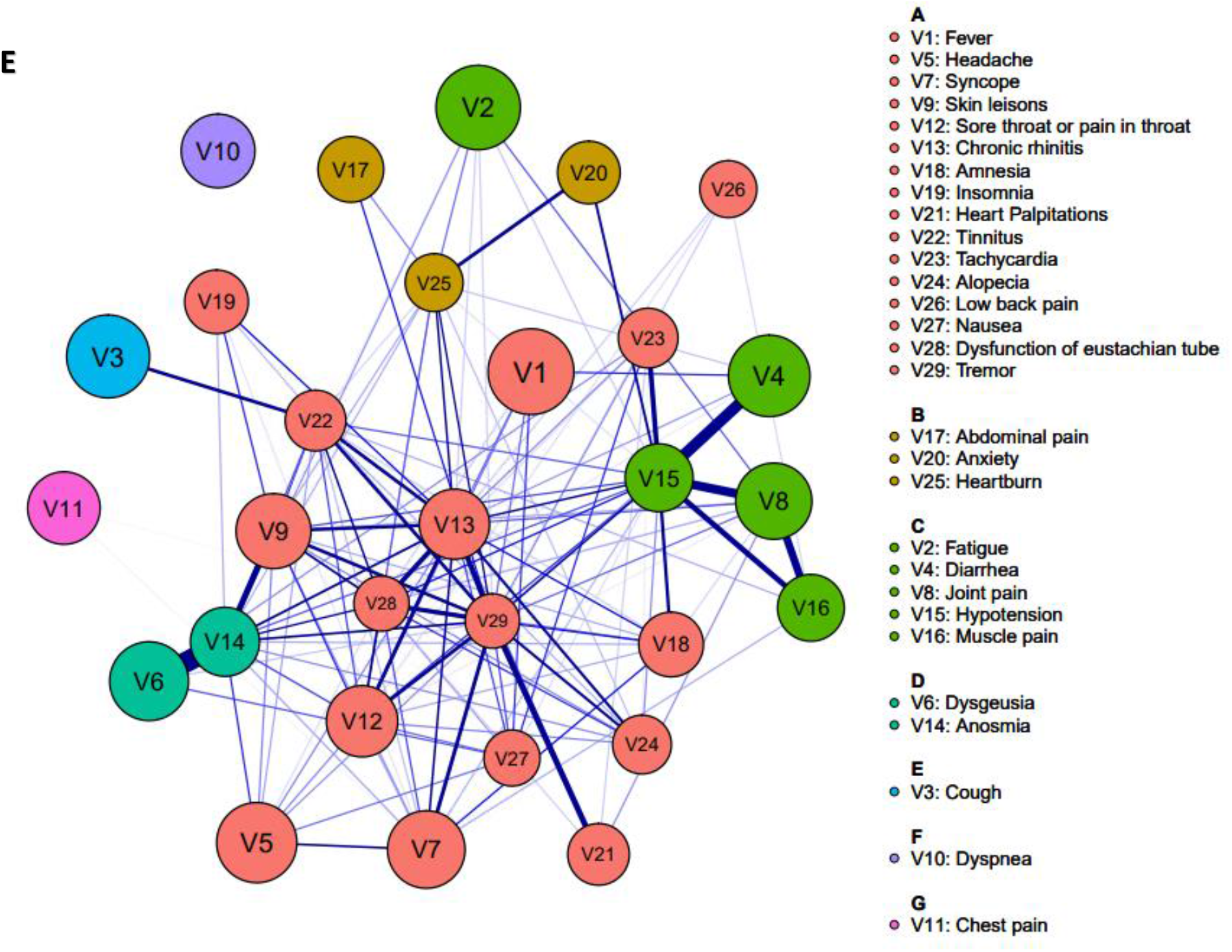
Demographics and symptom prevalence among SARS CoV-2 infected community dwellers at day 61+. (A) Bar and pie graphs showing distribution of age, ethnicity, and sex demographics (N=379) at days 61+. (B) Bar graph showing prevalence of symptoms reported at days 61+. (C) Graph of symptom clusters with (D) corresponding bar graphs demonstrating symptom ranking within each cluster, and (E) symptom network analysis showing relationship between each reported symptom. Each symptom is denoted as a node and the darker the line connecting symptoms indicates a stronger relationship.

### Symptoms at Day 0-10 and Predictive Value for Presence of Persistent Symptoms and Specific Symptom Cluster

At days 0-10 variables including age, sex, presence of symptoms, among others provided insight as potential predictors for becoming a long-hauler (Figure 3). Positive predictors included asymptomatic presentation, heart palpitations, chronic rhinitis, among others. In Figure 4, we evaluated which factors at days 0-10 would most likely result in grouping within one the five symptom clusters identified among long-haulers. The greatest magnitude between a feature and membership within a cluster included initial presentation with insomnia and chest pain-cough cluster, anxiety and the anxiety-tachycardia cluster, and nausea with the abdominal pain-nausea cluster. Non-modifiable factors such as age, sex, and ethnicity did not appear to be associated with belonging to a specific symptom cluster.

**Figure 3.**
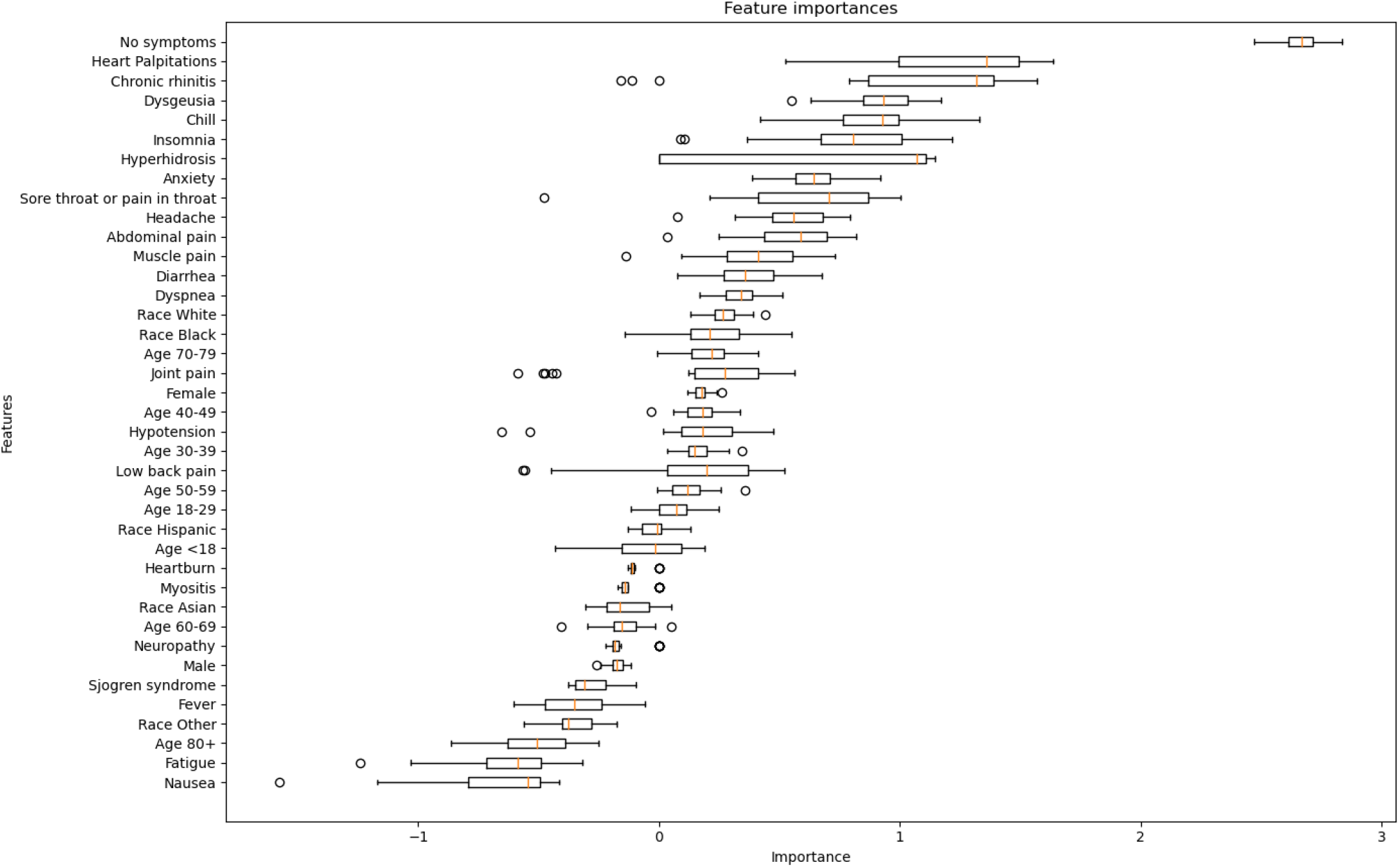
Key features during days 0-10 and their potential as indicators for developing prolonged COVID-19 symptoms or being a long-hauler. Bar graph showing factors that positively or negatively affect the probability of developing persistent symptoms among COVID+ community dwellers.

**Figure 4.**
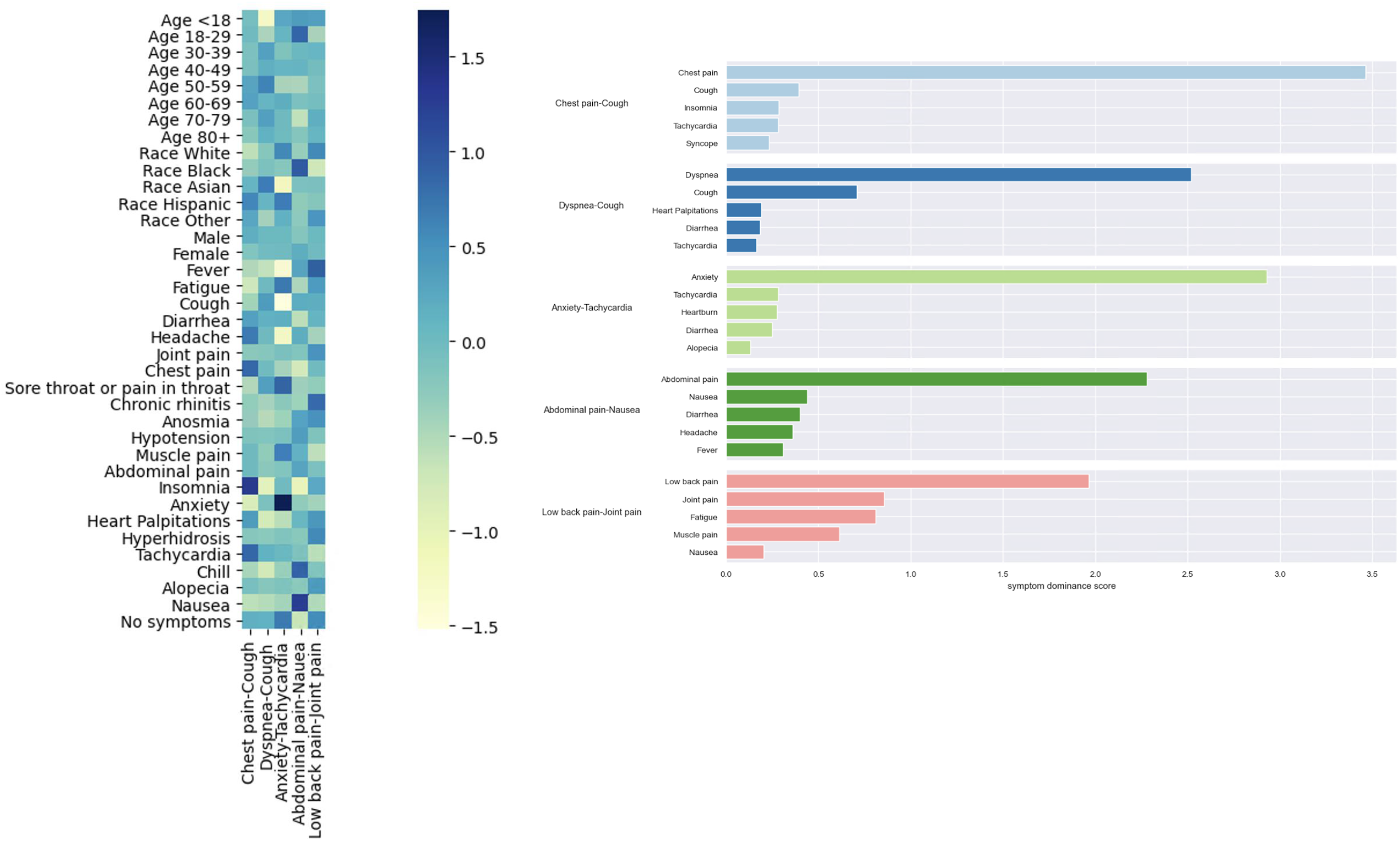
Presence of key indicators at days 0-10 predict inclusion into specific symptom clusters reported at day 61+. (A) NFM determined symptom clusters from SARS-CoV-2 infected community dwellers at day 61+; symptom clusters are named based upon the two most prevalent symptoms reported within each cluster. (B) Heat map demonstrating magnitude of association between key predictors from day 0-10 and assignment to a cluster with darker coloring indicating greater positive magnitude.

## Discussion

The current study provides much needed insight into early factors predisposing individuals for becoming long-haulers. These novel findings warrant additional investigations, discussion, and context within current knowledge about long-haulers. In reviewing our findings, we believe there are three key take-home points from our analyses.

The UC CORDS data set provides both patient-reported and clinician documented symptoms from SARS-CoV-2 infected patients. These symptoms are reported and recorded in real time which minimizes retrospective recall that has been used in the limited studies to date (4, 9). A few other important strengths from using the UC CORDS data set is that we exclude symptoms reported prior to SARS-CoV-2 infection to increase confidence in symptoms being attributable to becoming a long-hauler. The use of the data set allowed for a broad swath of symptoms, rather than being limited to a narrowly focused checklist of symptoms, which allows for a more sophisticated understanding of symptoms among long-haulers.

First, our observations suggest a developing picture of long-haulers potentially reflecting that Caucasian race, female sex, and normal BMI as common features specific to a sub-set of long-haulers. Although similar descriptions have been provided in other investigations (10) and the lay media, further corroboration is warranted. We observed a near normal distribution of age among long-haulers (Figure 2A), including those under the age of 18—with the mean age at 9.29 years. Although our study supported a potential association with female sex and higher likelihood of becoming a long-hauler, race appeared to be less predictive for both Caucasian and Hispanic ethnicity.

There has been conflicting information regarding whether asymptomatic individuals go on to become long-haulers, and roughly 32% of those reporting symptoms at day 61+ in our study were initially asymptomatic at the time of SARS-CoV-2 testing. Age distribution of all SARS-CoV-2 infected individuals at day 0-11 very closely mimicked that of the long-haulers, suggesting the latter group are distributed across all age groups with persons ages 50-59 range (± 20 years) representing more than 72% of the long-hauler population.

Secondly, the symptom experience among those who become long-haulers changes over time. Data from multiple studies converge to illustrate that many hospitalized and non-hospitalized survivors of COVID-19 experience persistent symptoms (10-16). The reported incidence of persistent symptoms varies; however, in the current study we report that 27% of community dwellers reported symptoms after 60 days. Some of the variability in symptom reporting and symptom association with long-haulers may be due to limitations inherent in rapid screening questionnaires in as much as these questionnaires inquire about symptoms that predominantly impact those with severe disease. Also, questionnaires may fail to inquire about emerging symptoms such as cognitive dysfunction (including “brain fog”), limiting the ability to accurately document such symptoms. Asymptomatic individuals may be less often intensely monitored due to an inherent notion of low risk for severe acute disease; however, this is problematic as asymptomatic individuals account for 32% of the long-haulers observed in this study. The symptom clusters observed among long-haulers vary compared to those at initial presentation. The evolution of these clusters may provide insight into the etiology of long-haulers in which elucidating sites of evolving tissue damage, and alterations in innate and adaptive immune inflammatory pathways might provide clarity in understanding the underlying pathophysiology.

In October 2020, the Tony Blair Institute for Global Change identified key characteristics among long-haulers, specifically that women appear to be at greater risk and those who are of working age (mean of age 45) (17). Our data align with these observations. Therefore, to our third key point, we observed that all ethnicities were affected as well as individuals who were initially asymptomatic. However, our use of ethnicity is limited to broad groups and lacks needed specificity, a limitation imposed by how data are recorded in the EHR. We therefore also assessed the most recently recorded BMI among long-haulers and those who had recovered from COVID-19 (Supplemental Figure 1). Mean BMI among long-haulers (by age group) ranged from 26 to 33, this may be due to limitations in our inclusion criteria (e.g. 5-year history with UC). Larger population-based studies will be needed to confirm and expand upon these observations. Undertaking detailed immune-profiling through emerging technologies such as the -omics platforms may identify key host phenotypes associated with the symptom clusters that we have described. We hope this article will prompt the development and implementation of longitudinal prospective studies that garner patient-generated reports of symptoms, rather than patient responses to questions generated by researchers -- this latter approach inherently constrained the answers we obtained. With such a new phenomenon, an ethnographic approach that focuses on understanding patients’ experiences would add an important lens to our analyses.

## Conclusion

Data are emerging to suggest that infection with SARS-CoV-2 may lead to prolonged and persistent symptoms. These long-term consequences of becoming a long-hauler are unclear, and further research is urgently needed to corroborate our findings. These findings include identifying a cohort of long-haulers with non-modifiable risk factors, which may have predicted the likelihood of persistent symptoms and/or assignment within given symptom clusters. Further research is needed to understand the underlying pathophysiology including host phenotypes associated with aberrant innate and adaptive immune responses following SARS-CoV-2 infection.

## Data Availability

Supplemental files are forthcoming for our lexicon for symptoms.

## Acknowledgement

*The dataset used in this study, University of California COvid Research Data Set (UC CORDS), is made available by the University of California Office of the President and the University of California Biomedical, Research, Acceleration, Integration, and Development. Biomedical computing facilities are supported by the National Center for Research Resources and the National Center for Advancing Translational Sciences, National Institutes of Health, through Grant UL1 TR001414. The content is solely the responsibility of the authors and does not necessarily represent the official views of the NIH*.

---

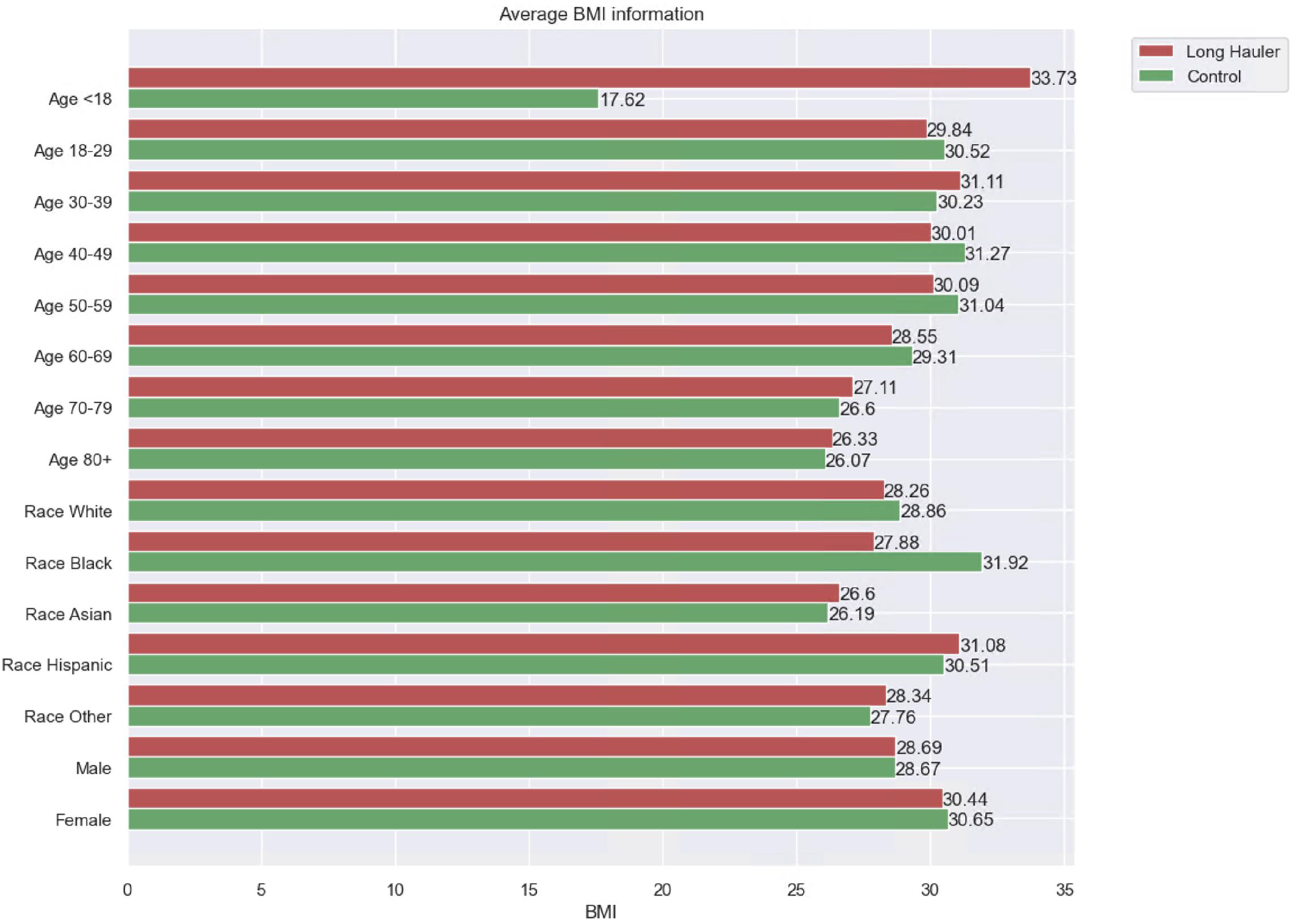

## References

1. COVID-19 Dashboard by the Center for Systems Science and Engineering [Internet]. [cited 2/10/2020]. Available from: https://gisanddata.maps.arcgis.com/apps/opsdashboard/index.html#/bda7594740fd40299423467b48e9ecf6.

2. COVID-19 Laboratory Confirmed Hospitalizations [Internet]. [cited 2/10/2020]. Available from: https://gis.cdc.gov/grasp/covidnet/COVID19_5.html).

3. Rubin R. As Their Numbers Grow, COVID-19 “Long Haulers” Stump Experts. JAMA. 2020;324(14):1381–3.

4. Carfì A, Bernabei R, Landi F, Gemelli Against COVID-19 Post-Acute Care Study Group. Persistent Symptoms in Patients After Acute COVID-19. JAMA. 2020 Aug 11;324(6):603–5.

5. Greenhalgh T, Knight M, A’Court C, Buxton M, Husain L. Management of post-acute covid- 19 in primary care. BMJ. 2020;370:m3026.

6. Lee DD, Seung HS. Learning the parts of objects by non-negative matrix factorization. Nature. 1999 Oct 21;401(6755):788–91.

7. Friedman J, Hastie T, Tibshirani R. Sparse inverse covariance estimation with the graphical lasso. Biostatistics (Oxford, England); Biostatistics. 2008;9(3):432–41.

8. Pons P, Latapy M. Computing communities in large networks using random walks (long version).. 2005.

9. Carvalho-Schneider C, Laurent E, Lemaignen A, Beaufils E, Bourbao-Tournois C, Laribi S, et al. Follow-up of adults with noncritical COVID-19 two months after symptom onset. Clin Microbiol Infect. 2020 Oct 5.

10. Huang C, Huang L, Wang Y, Li X, Ren L, Gu X, et al. 6-month consequences of COVID-19 in patients discharged from hospital: a cohort study. Lancet. 2021 Jan 16;397(10270):220–32.

11. Chopra V, Flanders SA, O’Malley M, Malani AN, Prescott HC. Sixty-Day Outcomes Among Patients Hospitalized With COVID-19. Ann Intern Med. 2020 Nov 11.

12. Goërtz YMJ, Van Herck M, Delbressine JM, Vaes AW, Meys R, Machado FVC, et al. Persistent symptoms 3 months after a SARS-CoV-2 infection: the post-COVID-19 syndrome? ERJ Open Res. 2020 Oct 26;6(4):00542,2020. eCollection 2020 Oct.

13. Davis HE, Assaf GS, McCorkell L, Wei H, Low RJ, Re’em Y, et al. Characterizing Long COVID in an International Cohort: 7 Months of Symptoms and Their Impact. medRxiv. 2020:2020.12.24.20248802.

14. Halpin SJ, McIvor C, Whyatt G, Adams A, Harvey O, McLean L, et al. Postdischarge symptoms and rehabilitation needs in survivors of COVID-19 infection: A cross-sectional evaluation. J Med Virol. 2021 Feb;93(2):1013–22.

15. Mandal S, Barnett J, Brill SE, Brown JS, Denneny EK, Hare SS, et al. ‘Long-COVID’: a cross-sectional study of persisting symptoms, biomarker and imaging abnormalities following hospitalisation for COVID-19. Thorax. 2020 Nov 10.

16. Meys R, Delbressine JM, Goërtz YMJ, Vaes AW, Machado FVC, Van Herck M, et al. Generic and Respiratory-Specific Quality of Life in Non-Hospitalized Patients with COVID-19. J Clin Med. 2020 Dec 9;9(12):3993. doi: 10.3390/jcm9123993.

17. Daniel Sleat, Ryan Wain, Brianna Miller. Long Covid: Reviewing the Science and Assessing the Risk. 2020 October 5,.

